# A Novel Fasting Mimetic (Mimio™) Improves Hunger, Digestion, Oxidative Stress, and Cardiometabolic Markers in Overweight Adults with Elevated HbA1c: a Double-Blind, Randomized, Placebo-Controlled Trial

**DOI:** 10.1101/2025.04.30.25326755

**Authors:** Azure D. Grant, Marie Crisel B. Erfe, Armenouhi Kazaryan, Paige L. Oliver, Jordan Moos, Veronica Luna, Noah Craft, Christopher H. Rhodes

## Abstract

This decentralized, double-blind, randomized, placebo-controlled trial investigated the impact of a novel fasting mimetic “Mimio*™*” on hunger, satiety, digestive symptoms, metabolism, cognition and wellbeing in overweight older adults with elevated HbA1c. Participants collected 2 weeks of baseline subjective data along with a fasted metabolic blood panel. The Mimio fasting mimetic, containing spermidine, nicotinamide, palmitoylethanolamide (PEA) and oleoethanolamide (OEA), or a placebo capsule was then taken before the first meal of the day for 8 weeks. Subjective measures were repeatedly collected throughout the study and metabolic bloodwork was repeated at the end of 8 weeks. 42 Participants were evaluated (47.6% female, BMI 27.6 (0.2) kg/m^2^, aged 62 ± 4, HbA1c 6.0 ± 0.1, n=23 intervention and n=19 placebo). Mimio improved more over time in all hunger and satiety metrics than placebo (Hunger and Satiety Composite Score Mann-Kendall p=2.2*10^-16^). More participants in the Mimio group improved daily ratings of hunger and appetite compared to placebo, including 91% vs. 47% of participants improving mealtime appetite across the study (Fisher’s Exact Test p=0.003). The Mimio cohort reported significantly less abdominal pain and bloating than the placebo group (Mann-Whitney U p<0.05). Mimio significantly reduced total cholesterol, LDL cholesterol, LDL particle number, oxidized LDL, non-HDL cholesterol and fasting glucose compared to placebo (Mann-Whitney U p<0.05). Changes in quality of life, three-factor eating questionnaire and cognitive failures did not differ significantly between Mimio and placebo. There were no significant differences in any adverse effects between Mimio and placebo. Mimio*™*, a daily fasting mimetic supplement, improves daily hunger and satiety, reduces oxidative stress, symptoms of indigestion and improves cardiometabolic health markers in overweight older adults with elevated HbA1c. This is the first study to show that fasting mimetic supplementation can recreate clinical fasting-like cardiometabolic benefits without lifestyle changes or the need to fast.

**Trial Registration:** The trial was IRB approved and registered with ClinicalTrials.gov (Pro00080269).

## Introduction

Intermittent fasting has gained significant scientific and clinical interest in recent years due to its well-documented beneficial effects on conditions ranging from cardiovascular disease to metabolic disorders, neurodegenerative conditions, cancer and autoimmune disease^1–10^. Moreover, prolonged periods of fasting remain one of the only reliable methods of extending lifespan in model organisms due their direct modulation of the cellular aging process^1–4^. Unfortunately, fasting’s long term impracticality and safety concerns limit its widespread adoption. Despite its potential to significantly improve human health by helping to treat, prevent, or delay disease and enhance longevity, the duration of fasting required to achieve such benefits (typically greater than 24 h)^1–3^ is a significant burden to quality of life. Moreover, this fasting duration is often infeasible or dangerous for numerous populations, including adolescents^11^, endurance athletes^12^, lean or underweight young people^13^, the elderly, those with a history of eating disorders and pregnant or breast-feeding women^11^. Interventions capable of mimicking the beneficial cellular, metabolic, immune, and longevity-enhancing effects of fasting without the need to abstain from food hold great potential therapeutic value for both the treatment and prevention of chronic health conditions, especially for those for whom fasting may be contraindicated.

Spermidine, nicotinamide, palmitoylethanolamide (PEA) and oleoylethanolamide (OEA) are endogenous human metabolites that appear to improve clinical markers of disease, extend lifespan and possess both anti-inflammatory and antioxidant properties^14–17^. A previous study conducted by the Zivkovic Lab at UC Davis identified higher plasma concentrations of these metabolites after a 36 h fast compared to a 12 h overnight fast and demonstrated that a combination of spermidine, 1-methlynicotinamide (1-MNA, a cellular derivative of nicotinamide), PEA and OEA could mimic many of the functional benefits of prolonged fasting including significant anti-inflammatory and antioxidant effects *in vitro* and extending lifespan by 96% in *C. elegans*^18^. This fasting mimetic formulation has also been shown in a pilot clinical study to be orally bioavailable, well-tolerated, and capable of replicating anti-inflammatory, antioxidant and cardioprotective effects similar to fasting when taken in the postprandial state^19^.

The present study investigated the impacts of this novel fasting mimetic formulation (Mimio Biomimetic Cell Care, “Mimio”) on subjective and objective measures of metabolic and digestive health in overweight adults with elevated hemoglobin A1c (HbA1c) in a randomized, double-blind, placebo-controlled setting. Subjective measures of hunger and satiety, gastrointestinal symptoms, cognitive failures and quality of life were administered. Additionally, blood samples for measurement of NMR lipoprofile, oxidized LDL, hsCRP, HbA1c, insulin and plasma glucose were collected before and after intervention.

## Methods

### Eligibility

Participants needed to be at least 55 years of age, have a BMI between 25 and 29.9 and an HbA1c level of at least 6.0. Female participants were required to be menopausal (>12 months since last menstrual period) and not use exogenous hormones. If taking any fiber supplements, prescription medication for cholesterol (e.g., statins) or other class of medications for metabolic disorders such as metformin, ACE inhibitors, beta blockers, rapamycin, thiazolidinediones, etc., participants were required to maintain a stable dose for at least 4 weeks prior to randomization and throughout the course of the study.

If taking any other supplement products such as fish oil, resveratrol, alpha-ketoglutarate, berberine, ashwagandha, pre- and probiotics, melatonin, NAD+ precursors (e.g., niacin, nicotinamide riboside (NR), nicotinamide mononucleotide (NMN), NADH), fisetin, astaxanthin, urolithin-A, CoQ-10, green tea extract/EGG, or pterostilbene, participants were required to stop from at least 1 week prior to randomization through the end of the study. Participants were required to abstain from cannabis products and maintain a consistent diet throughout the study.

Participants could not be taking any GLP-1 agonists or incretin mimetics, any investigational therapies, or be regular users of over-the-counter allergy or pain medications (i.e., >3-4 times per week). Participants could not be diagnosed with diabetes, eating disorders, any substance abuse, anemia, CVD, or cancer. Finally, participants could not have any known hypersensitivity or previous allergic reaction to nicotinamide, palmitoylethanolamide, oleoylethanolamide, spermidine.

Participants needed to be able to read and understand English, use a personal smartphone device and download the mobile application Chloe by People Science, receive shipment of the product at an address within the United States and be willing travel to a local Quest Diagnostics laboratory (Quest Diagnostics, Secaucus, NJ, USA) for fasted blood sample collection twice during the course of the study.

### Study Design

Participants completed up to a 14-week study consisting of a screening period, randomization and shipping period, a 2-week baseline period and an 8-week product or placebo use period. A schedule of all study activities is provided in **Table 1**.

**Table 1.**
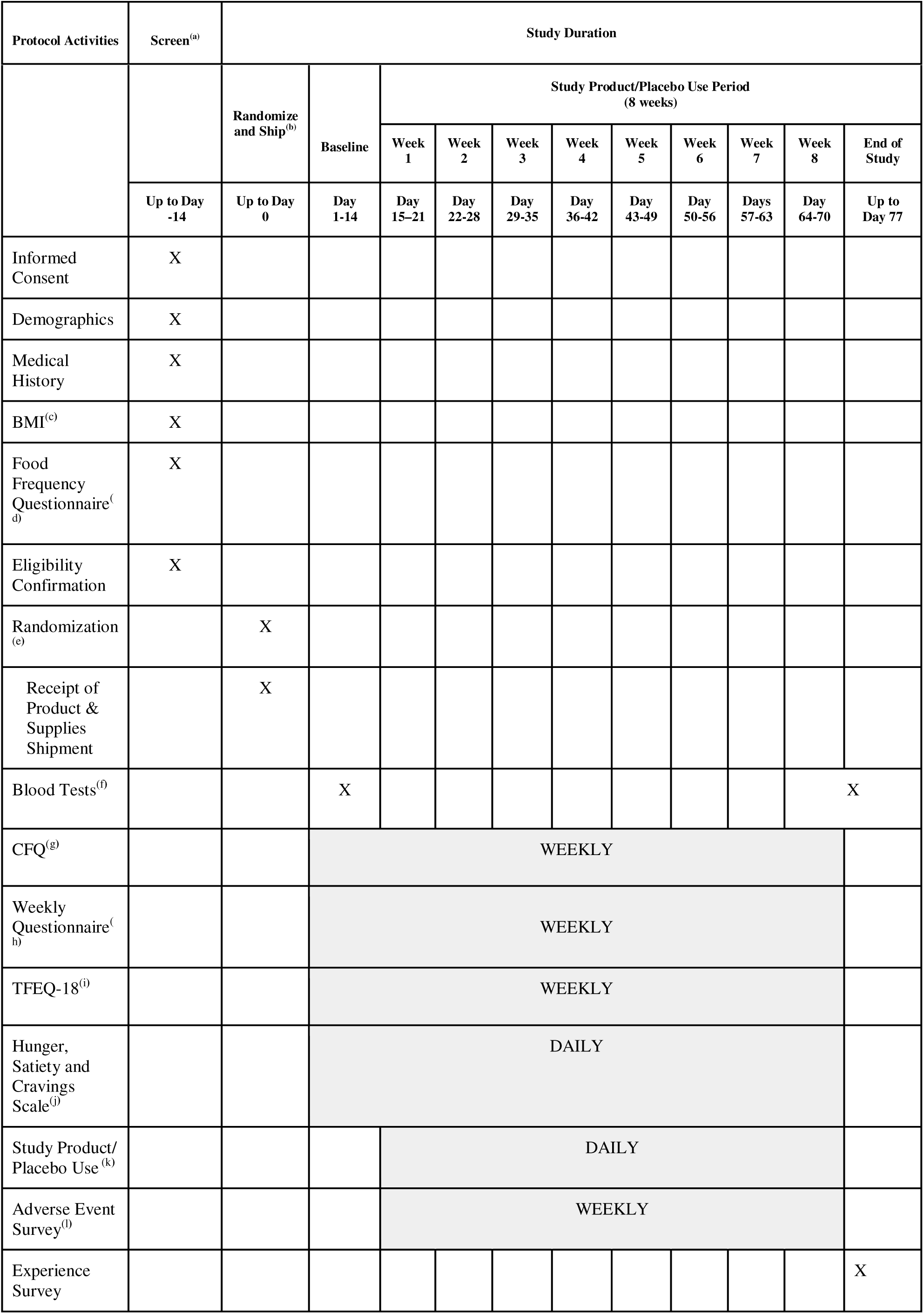
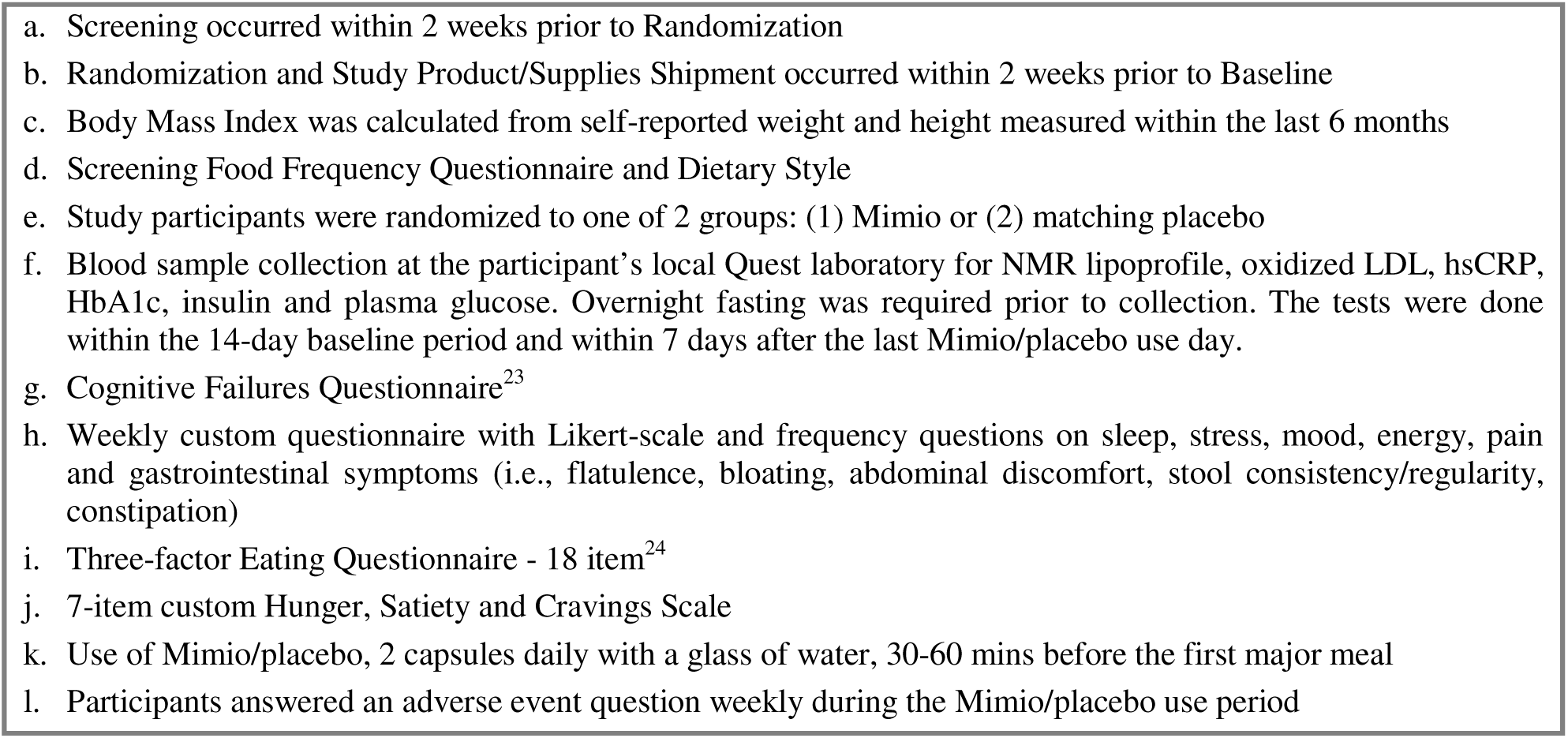
Study Activities.

Participants were randomized to one of 2 groups by AK: Mimio capsules or matching placebo capsules, with stratification to ensure even balance of male and female participants in each group. The investigators, study team and participants were blinded to the group assignment. Participants received the study product and study supplies after randomization. Demographic, medical history and concomitant medications data were collected during the screening period in Chloe. All assessments and surveys were administered in the Chloe mobile app. Blood sample collection occurred at the participant’s local Quest laboratory. Adverse events were both actively, via a weekly prompt, and passively collected throughout the study.

### Recruitment and Consent

Participants who had never tried Mimio were recruited through the People Science community and social media channels. Virtual electronic informed consent, including a study-specific privacy authorization and the California Experimental Subject’s Bill of Rights (as applicable) were provided through Chloe. A digital copy of the signed consent was accessible by the participant through their Chloe profile. Eligible participants who provided virtual electronic consent were enrolled into the study.

### Mimio Supplement and Placebo

Each dose of Mimio, to be taken 30-60 minutes before the first major meal of the day, contained Nicotinamide (250mg), Ultra-Micronized Palmitoylethanolamide (600mg), Oleoylethanolamide (400mg), Spermidine (8mg); Hydroxypropyl-Methylcellulose (capsule), Silicon Dioxide and Magnesium Stearate. The placebo capsules contained Micro Crystalline Cellulose (250mg), Hypromellose and Titanium Dioxide.

### Study Platform

All study tasks, communication and data were collected and assembled via the web and mobile-app based data collection platform, the Consumer Health Learning and Organizing Ecosystem (Chloe) by People Science (People Science Inc., Los Angeles, CA, USA). Hence, participants were able to collect data from their homes or on the go. All data was securely stored on People Science Amazon Web Services HIPAA-compliant servers. The Chloe platform contains modules for building and managing surveys, landing pages, marketing outreach with tracking tools for recruitment, audited electronic consent forms and electronic case report forms (eCRF), data management and analytics using an integrated relational database. Data from completed assessments was automatically collected for analysis. Study monitoring was conducted using reporting features.

### Subjective Measures

The study’s primary outcome was a daily Hunger, Satiety and Cravings Scale consisting of 7 items on a 0-5 scale (strongly disagree to strongly agree). Questions pertained to hunger, appetite, cravings and satiety in the past 24 hours (e.g., “yesterday I craved foods I think of as unhealthy,” “yesterday I felt satisfied after my meals, neither too full nor hungry”). This daily survey began at baseline and was administered throughout the product/placebo use period. Digestive symptoms and quality of life were evaluated using a weekly survey. The weekly survey consisted of a 13-item set of Likert scales relating to sleep, stress, mood, energy, food noise and gastrointestinal symptoms (i.e., flatulence, bloating, abdominal discomfort, nausea and bowel movement regularity). The cognitive aspect of eating behavior was evaluated using weekly repetitions of the Three-Factor Eating Questionnaire (TFEQ-18). The TFEQ-18 is a validated, 18-item questionnaire representing the derived factors of Cognitive Restraint, Uncontrolled Eating and Emotional Eating^20^. See **Table 1** for schedule of assessments.

As fasting may be associated with cognitive function^21^, The Cognitive Failures Questionnaire (CFQ) was administered at baseline and weekly across the product and placebo use period. The CFQ is designed to assess a person’s self-rated proneness to committing cognitive slips and errors of perception, memory, motor functioning and absent-mindedness in the completion of everyday tasks^22,23^. Finally, to further evaluate product safety, participants were asked about any negative experiences at the end of each week and filled out a study experience survey after protocol completion.

### Blood Measures

Blood was drawn after at least an 8 h overnight fast at baseline and end of study for evaluation of the following markers: plasma glucose (mg/dL), insulin (uIU/mL), hemoglobin A1c (HbA1c, % of total hemoglobin), hs-CRP (mg/L), triglycerides (mg/dL), total cholesterol (mg/dL), HDL cholesterol (HDL-c, mg/dL), calculated LDL cholesterol (LDL-c, mg/dL), total cholesterol/HDL-c ratio, calculated non-HDL Cholesterol (mg/dL) and triglyceride/HDL-c ratio. An NMR lipid profile provided the following additional lipid markers: LDL particle concentration (LDL-p, nmol/L), small LDL-p concentration (nmol/L), LDL size (nm), HDL particle concentration (HDL-p, µmol/L), large HDL-p concentration (µmol/L), HDL size (nm), large VLDL particle concentration (VLDL-p, nmol/L), VLDL size (nm) and oxidized LDL (OxLDL, U/L). Samples were collected and analyzed by Quest Diagnostics.

### Sample, Data Evaluability and Statistics

Sample size was determined based on estimated change in the Hunger and Satiety scale. Data analysis was conducted in Python (version 3.9.13) Jupyter Notebooks (Jupyter LLC, NYC, NY, USA). Participants were considered evaluable for analysis if they completed 8 weeks of Mimio/placebo use, baseline measurements and at least 70% of their questionnaires. Missing data in daily surveys were linearly interpolated and individual days in which a dose of Mimio was missed were omitted. Data were tested for normality using the Shapiro-Wilk test.

The primary objective of this study was to evaluate the impact of Mimio on a daily scale of hunger & satiety. We aggregated individual changes in the survey from the baseline period to week 8 and compared placebo and Mimio groups using parametric statistics (t-tests or repeated measures ANOVA (rmANOVA) when data were normally distributed and Mann Whitney U ranksum tests when data were non-normally distributed. Mixed effects models were used to evaluate impact of demographic factors. Daily data were evaluated both on change in absolute scores and by within-individual change in scores via Mann-Kendall (MK) trend over time. Difference in percentage improvers assessed by Fisher’s Exact Tests (FET).

Secondary objectives, including change in bloodwork, cognitive failures questionnaire (CFQ), cognitive and behavioral components of eating (TFEQ-18) and change in self-reported sleep, stress, mood, energy, pain and gastrointestinal symptoms (i.e., flatulence, bloating, abdominal discomfort, stool consistency/ regularity, constipation), were evaluated using absolute before and after scores, by comparing within-individual magnitude and direction of change and proportion of individuals who improved in each group. FET was used to evaluate differences in the occurrence of categorical variables in the weekly wellness and digestive health surveys.

AEs were summarized by group, displaying number of participants in each group, severity, possible relation to the study intervention and resolution.

## Results

### Recruitment and Conduct

This study was approved by the Advarra Institutional Review Board (Pro00080269) and registered with clinicaltrials.gov (NCT06790407, 4/25/2024). All participants gave informed consent. All study procedures were conducted by People Science following Good Clinical Practice (GCP) guidelines and in accordance with the ethical principles of the Declaration of Helsinki and its amendments. Recruitment occurred 7/29/2024-10/7/2024. The trial concluded after recruited participants completed study activities. The full protocol can be accessed at clinicaltrials.gov.

### Demographics

A total of 42 individuals were included (Mimio n=23, Placebo n=19) for all primary and secondary outcomes except blood work, with an intervention adherence rate of 94%. A total of 33 participants were evaluated for blood markers with 9 participants being excluded based on violations to protocol compliance including failure to fast during blood draws or to complete bloodwork within a week of study completion. The mean age (SEM) was 62 ± 4 years. BMI mean (SEM) was overweight but not obese, 27.7 ± 0.16. The mean HbA1c level was 6.0 ± 0.1. Sex, age, ethnicity, BMI and baseline bloodwork did not differ by group. Fasting bloodwork values did not differ statistically for any metric at baseline between the Mimio and placebo groups. Demographics are described in **Table 2**. Laboratory values at baseline are presented in **Table 3** and are graphed in **Supplemental Figure 1**.

**Table 2.**
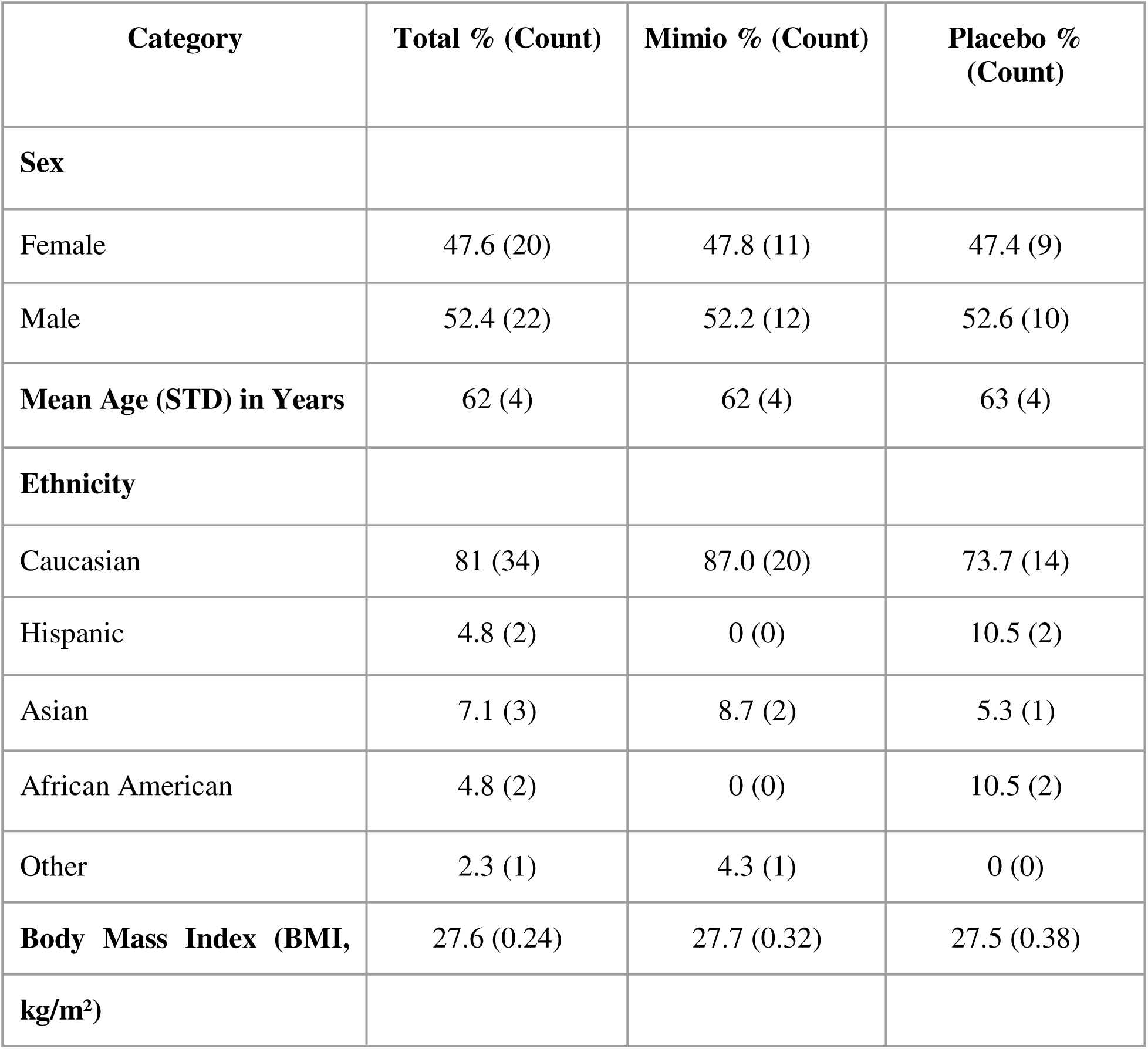
Demographics.

**Table 3.**
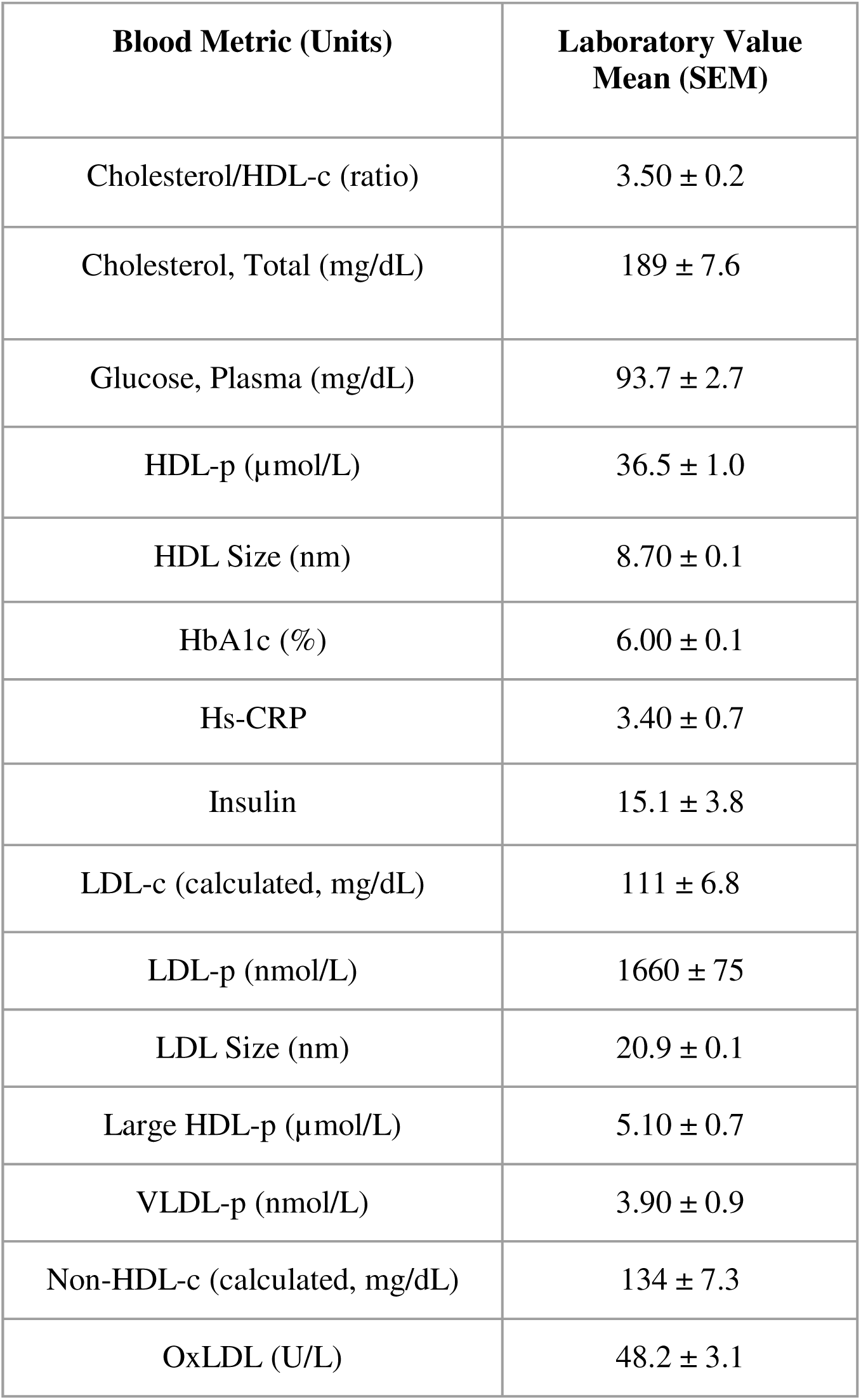
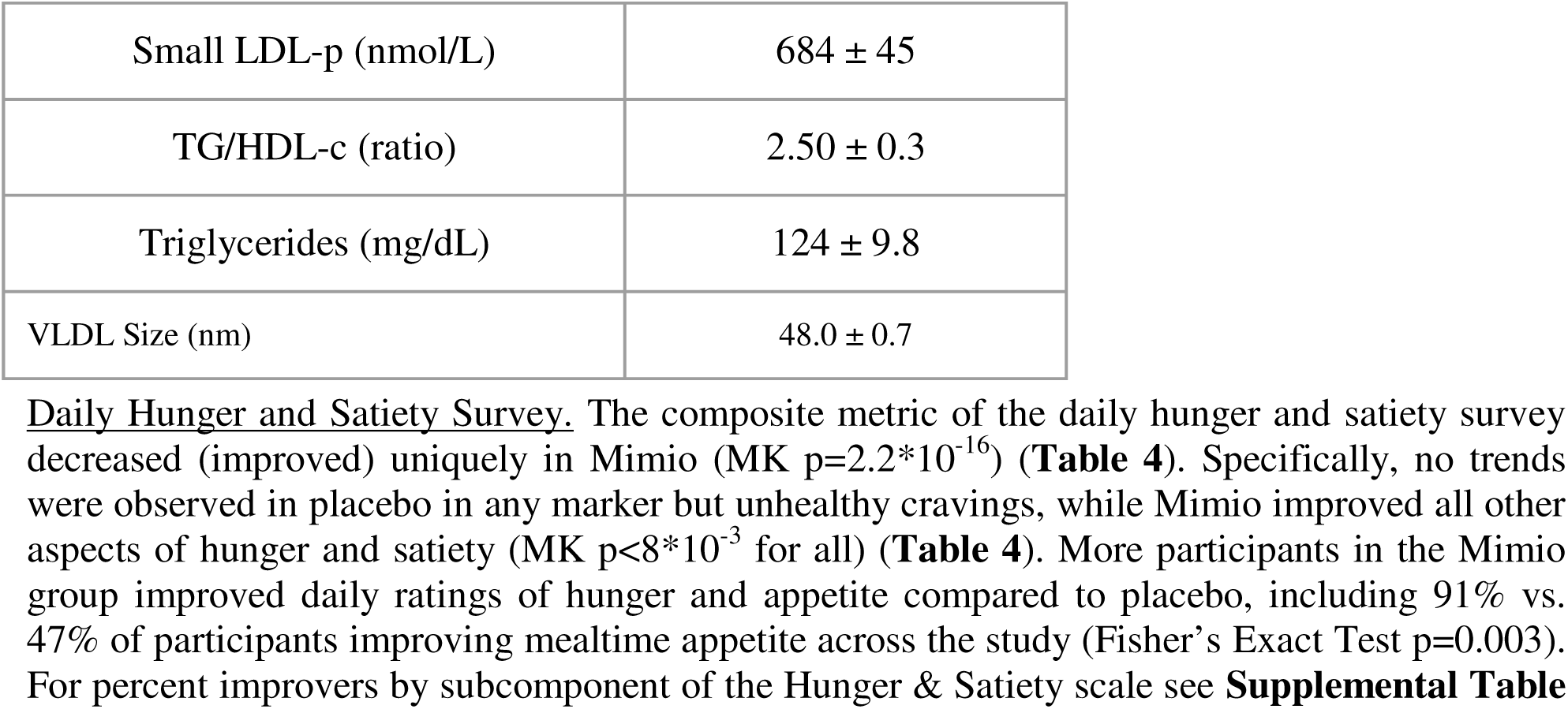
Baseline Metabolic Values.

| Blood Metric (Units) | Laboratory Value<br>Mean (SEM) |
| --- | --- |
| Cholesterol/HDL-c (ratio) | 3.50 ± 0.2 |
| Cholesterol, Total (mg/dL) | 189 ± 7.6 |
| Glucose, Plasma (mg/dL) | 93.7 ± 2.7 |
| HDL-p (μmol/L) | 36.5 ± 1.0 |
| HDL Size (nm) | 8.70 ± 0.1 |
| HbA1c (%) | 6.00 ± 0.1 |
| Hs-CRP | 3.40 ± 0.7 |
| Insulin | 15.1 ± 3.8 |
| LDL-c (calculated, mg/dL) | 111 ± 6.8 |
| LDL-p (nmol/L) | 1660 ± 75 |
| LDL Size (nm) | 20.9 ± 0.1 |
| Large HDL-p (μmol/L) | 5.10 ± 0.7 |
| VLDL-p (nmol/L) | 3.90 ± 0.9 |
| Non-HDL-c (calculated, mg/dL) | 134 ± 7.3 |
| OxLDL (U/L) | 48.2 ± 3.1 |
| Small LDL-p (nmol/L) | 684 ± 45 |
| TG/HDL-c (ratio) | 2.50 ± 0.3 |
| Triglycerides (mg/dL) | 124 ± 9.8 |
| VLDL Size (nm) | 48.0 ± 0.7 |

**Table 4.** Trends Over Time in Hunger & Satiety.

| Daily Metric | Mimio<br>Mean Trend (Mann-Kendall<br>p-value) | Placebo<br>Mean Trend (Mann-Kendall<br>p-value) |
| --- | --- | --- |
| Unhealthy Cravings | Decreasing ($p=3.8*10^{-9}$) | Decreasing ($p=1.4*10^{-2}$) |
| Eating Only When Hungry | Increasing ($p=7.1*10^{-3}$) | No Trend ($p>0.05$) |
| Distraction From Cravings | Decreasing ($p=7.7*10^{-9}$) | No Trend ($p>0.05$) |
| Postprandial Satiety | Increasing ($p=1.2*10^{-5}$) | No Trend ($p>0.05$) |
| Mealtime Appetite | Decreasing ($p=1.7*10^{-12}$) | No Trend ($p>0.05$) |
| Maximum Daily Hunger | Decreasing ($5.6*10^{-14}$) | No Trend ($p>0.05$) |
| Overall Daily Hunger | Decreasing ($2.5 \times 10^{-13}$) | No Trend ($p > 0.05$) |
| Composite Score | Decreasing ($2.2 \times 10^{-16}$) | No Trend ($p > 0.05$) |

**Figure 1.**
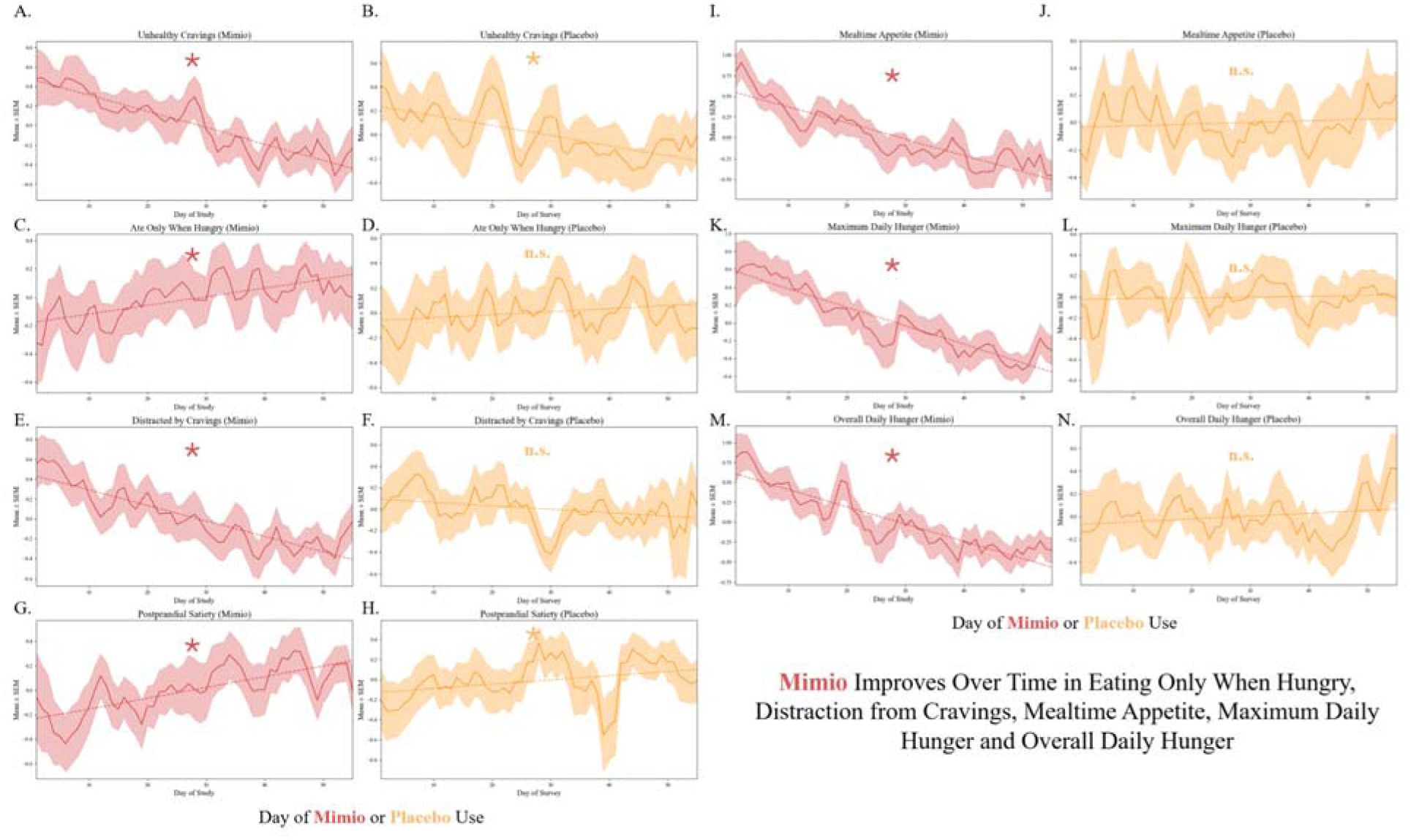
**Mimio is Uniquely Associated with Eating When Hungry, Reduced Cravings, Lower Mealtime Appetite and Less Hunger** People taking Mimio (red, left columns) exhibited a statistically significant Mann-Kendall trend over time to improvement unhealthy cravings (A), eating only when hungry (C), distraction from cravings (E), postprandial satiety (G), mealtime appetite (I), maximum daily hunger (K) and overall daily hunger (M) (MK trend over time p<0.05). The placebo group (yellow, right columns) statistically improved only unhealthy cravings (B) and postprandial satiety (H) (MK trend over time p<0.05).

### Weekly Digestive Symptom and Wellness Survey

Weekly measures did not differ statistically at baseline (Student’s t-tests p>0.05). Frequency of bloating was reduced by the final week of the study (Student’s t-test p=0.001), with 87.5.6% vs 38.1% of participants reporting no bloating in the final week **(Figure 2A)**. Additionally, abdominal pain frequency was reduced (Student’s t-test p=0.008), with 83.3% vs. 45.5% of participants reporting never (**Figure 2B**). Other digestive metrics trended toward improvement with Mimio (**Supplemental Figure 2)**. Weekly metrics of self-reported sleep duration, quality, anxiety, contentedness, low mood, food noise, stress and energy did not differ by group (**Supplemental Figure 3**).

**Figure 2.**
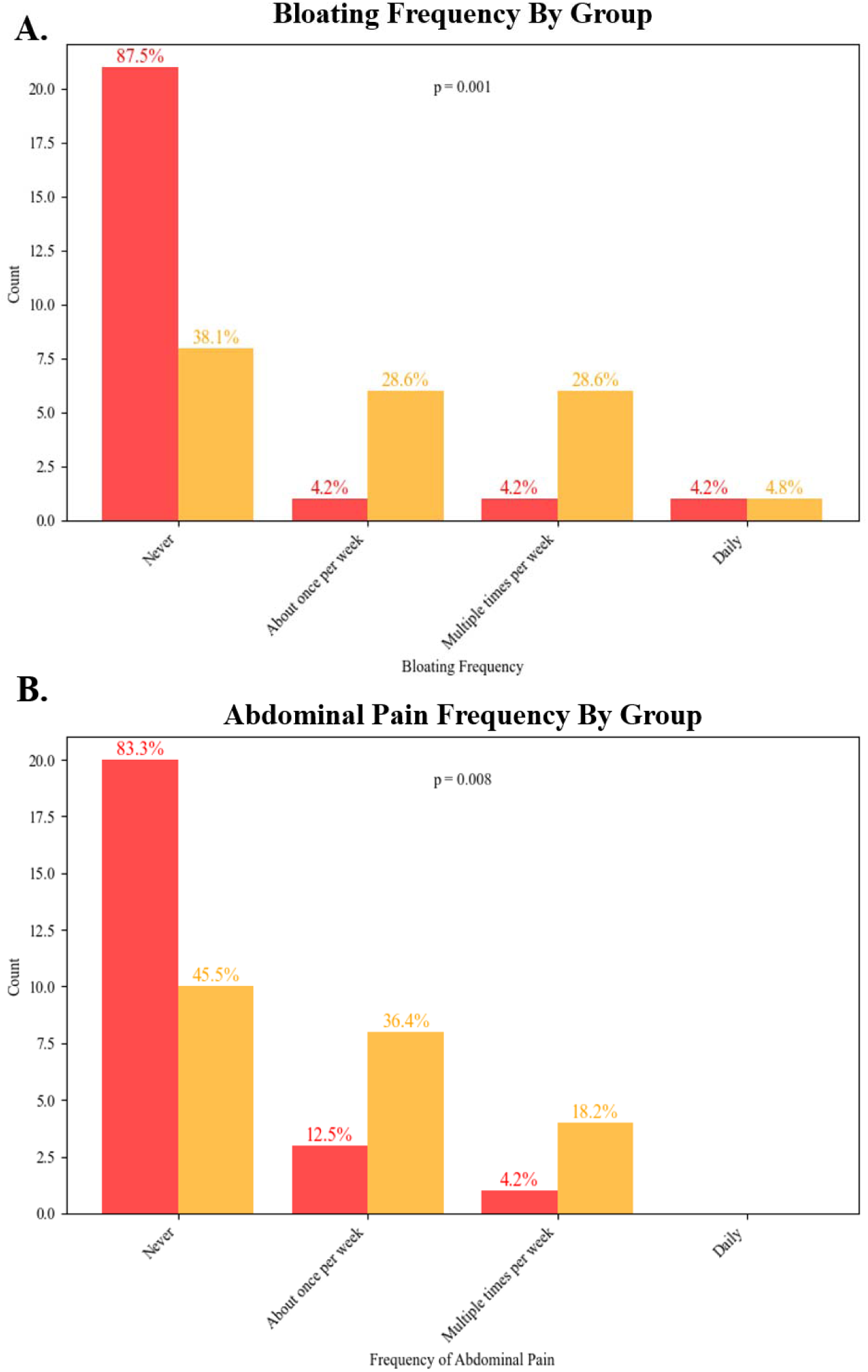
**Mimio Reduced Frequency of Bloating and Abdominal Pain Compared to Placebo** Mimio (red) reduced the frequency of bloating (A) and abdominal pain (B) compared to placebo (yellow). Data represent the final week of the study. Percentages indicate the proportion of that cohort that gave a given answer (e.g., 87.5% of Mimio participants “Never” experienced bloating in the final week of the study, vs. 38.1% of placebo participants). The y-axis of the histogram represents the count of individuals.

### Metabolic Bloodwork

Blood work analysis revealed several significant improvements in cardiometabolic risk factors and oxidative stress markers in the Mimio group compared to the placebo including reductions in total cholesterol, fasting glucose, LDL cholesterol, LDL particle number, non-HDL cholesterol, and Oxidized LDL (**Figure 3**). Of these, the most substantial were a 5.4% drop in LDL-p with Mimio vs. a 4.8% rise in placebo (student’s t-test p=0.008; net change vs placebo 10.2%) and an 8.6% drop in Oxidized LDL with Mimio vs. a 4.3% rise in placebo (student’s t test p=0.047; net change vs. placebo 12.9%). Statistical trends evaluated by raw change per metric and % within-individual change per metric were consistent across all metrics. There was no statistically significant change across other blood measures between the placebo and Mimio groups including insulin, CRP, HDL-c, HbA1c, or triglycerides. Complete values for each blood metric can be found before and after in **Supplemental Figure 1.** Groups did not differ at baseline in values or proportion of cohorts “In Range” (data not shown).

**Figure 3.**
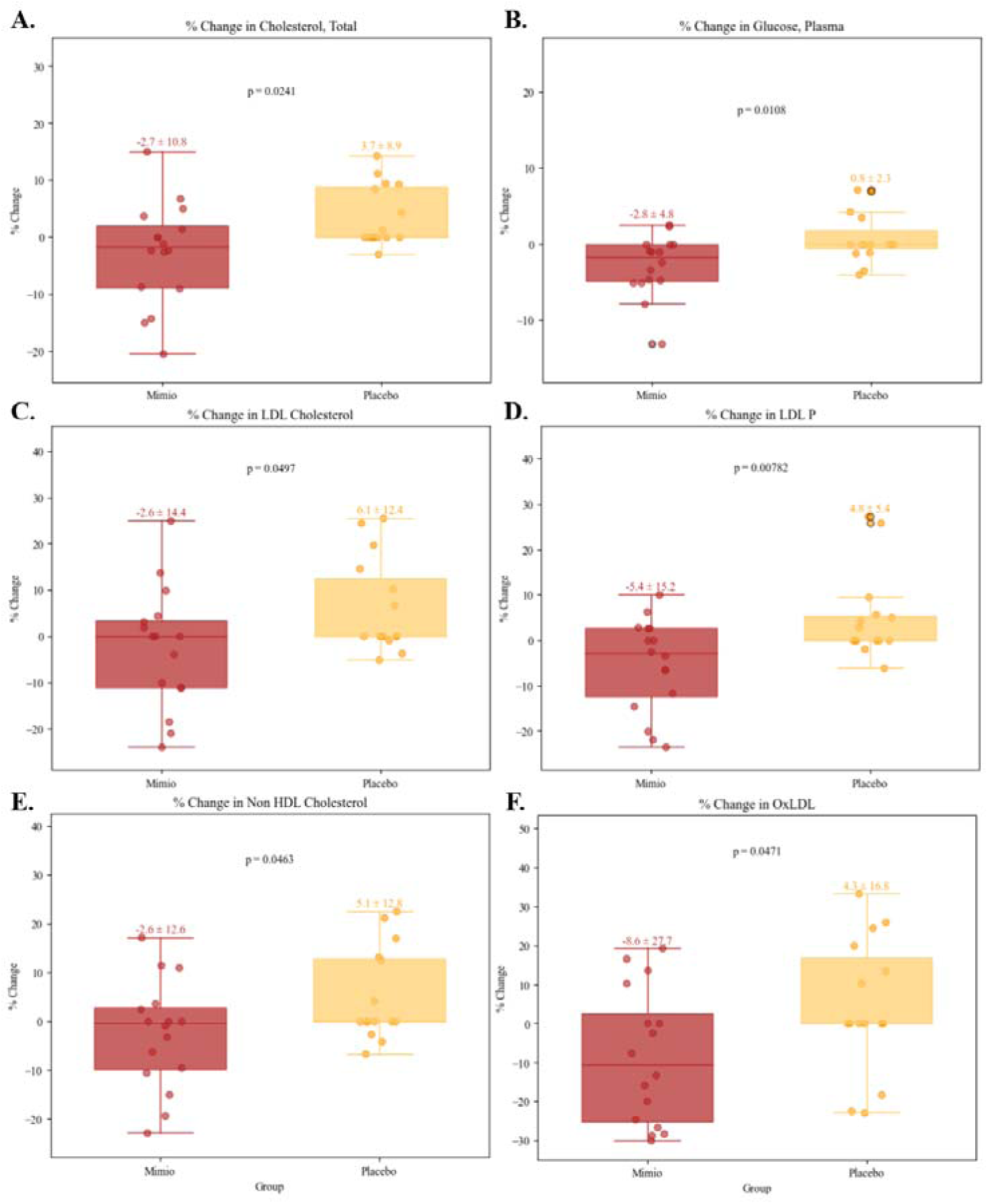
**Mimio Improves Total Cholesterol, Glucose, LDL-c, LDL-p, Non-HDL-c and Oxidized LDL Compared to Placebo** Mimio improves cardiometabolic and oxidative stress markers compared to placebo. Mimio was associated with a reduction in total cholesterol (A), glucose (B), LDL-c (C ), LDL-p (D), Non-HDL-c (E), and oxidized LDL (F). Mimio is shown in red, placebo in yellow. Values above box and whisker plots represent mean ± SEM of % change from baseline to end of study. Scattered dots represent individual participants’ values. P-values are displayed on each plot.

### Adverse Events

Adverse event rates did not differ statistically between the Mimio and placebo groups. Only 1 AE, a case of mild diarrhea, occurred during product use in the Mimio group and was considered possibly attributable to the intervention. By comparison, 5 digestive AEs occurred in the Placebo group. See **Supplemental Materials and Supplemental Table 2.**

### Three Factor Eating Questionnaire (TFEQ-18)

Mimio and Placebo groups did not differ in any TFEQ sub score: Cognitive Restraint, Uncontrolled Eating or Emotional Eating. Starting and ending values for each sub score are shown by group in **Table 5** and graphed in **Supplemental Figure 4-5**. Within-individual change from baseline to end of product use and did not vary by group (Mann-Kendall and rm-ANOVA p>0.05 for all subscales). Percent improvers did not vary by group (FET p>0.05 for all subscales, data not shown).

**Table 5.** TFEQ-18 Scores.

| Sub Score | Mimio Start | Mimio End | Placebo Start | Placebo End |
| --- | --- | --- | --- | --- |
| Cognitive Restraint | $2.47 \pm 0.12$ | $2.22 \pm 0.13$ | $2.56 \pm 0.13$ | $2.41 \pm 0.15$ |
| Uncontrolled Eating | $2.65 \pm 0.08$ | $2.70 \pm 0.10$ | $2.63 \pm 0.12$ | $2.63 \pm 0.15$ |
| Emotional Eating | $2.38 \pm 0.11$ | $2.00 \pm 0.10$ | $2.33 \pm 0.14$ | $2.01 \pm 0.13$ |

## Discussion and Conclusions

Prolonged fasting is one of the few interventions consistently shown to extend lifespan and health span in model organisms. Despite its robust potential in clinical applications to treat, prevent, or delay most major diseases, the safety concerns, impracticality and patient burden associated with fasting limit widespread adoption. As such, interventions that *mimic* the beneficial effects of fasting could provide a safe and accessible alternative, especially for those unable to regularly fast. In this randomized, double-blind, placebo-controlled trial we investigated the impact of a novel fasting mimetic (Mimio) on hunger, satiety, digestive symptoms, cardiometabolic health, cognition and wellbeing in overweight older adults with elevated HbA1c. Mimio induced changes in both the subjective experience of hunger, satiety and digestion as well as the cardiometabolic blood profile over 8 weeks of supplementation. Daily hunger and satiety metrics improved substantially compared to placebo, with 91% vs. 47% of participants improving appetite across the study and all metrics exhibiting a statistical trend in the direction of improvement. Supplementation with Mimio also resulted in significant reductions in self-reported abdominal pain and bloating compared to placebo. Most strikingly, Mimio also induced fasting-like improvements to cardiometabolic blood markers with LDL particle number, total cholesterol, oxidized LDL-c, LDL-c, nonHDL-c and glucose concentrations improved significantly compared to placebo. Together, daily use of Mimio appears safe and tolerable and produced significant improvements to multiple metrics of hunger control, indigestion, lipid composition, cholesterol metabolism, oxidative stress and glycemia. These effects successfully “mimic” the metabolic changes observed during prolonged fasting without changes to diet or lifestyle.

Elevated levels of LDL particles are associated with an increased risk of cardiovascular disease (CVD)^25,26^. Among these, the small dense particles are more prone to penetrate the arterial wall, become oxidized and contribute to atherosclerosis^27–29^. In some studies, reduction in small dense LDL particle concentration have been linked to decreased CVD risk, especially when accompanied by improvements in other lipid parameters. For instance, therapies that lower LDL particle number and increase particle size have been associated with reduced progression of coronary artery disease^28–30^. Similarly, the change in fasted glucose here corresponds to a few mg/dL on average or 5-6 mg/dL in the most responsive quartile. However, these changes occurred over a short period of time with no recommended changes to diet or exercise. Given the linear progress over time in subjective sensations of hunger and satiety in the Mimio cohort, with no evidence of a tolerance effect, it is possible that blood metrics would also improve further with prolonged usage of the fasting mimetic.

Together, as the present study provided only 8 weeks of intervention and did not strictly control for dietary intake, it is encouraging to see even modest improvements in lipid composition and fasting glucose. Longer duration studies will be necessary to determine the extent of these effect. The addition of more frequent lipid monitoring via point of care devices, continuous glucose monitoring or dietary standardization may clarify the linearity of improvement over time. Furthermore, unlike previous studies utilizing nicotinamide, spermidine, PEA and OEA individually, this study of the Mimio combination was performed in a healthy population with no diagnosed metabolic, immune, cardiovascular or cognitive conditions or diseases, which may also account for a more modest effect size than observed in previous studies on similar biomarkers. However, the ability for a combination of natural molecules to significantly improve multiple relevant cardiometabolic risk factors and robustly improve multiple measures of hunger control in an already healthy population in just 8 weeks without the need to change diet or lifestyle presents a promising new approach for health optimization and disease prevention.

The present study builds upon findings from pre-clinical and clinical studies of the individual impacts of nicotinamide/1-MNA, spermidine, PEA and OEA and their combination (Mimio) on feeding behavior and cardiometabolic markers. 1-MNA has been shown to extend lifespan in *C. elegans* through the induction of cellular antioxidant mechanisms^31^. Although there is no current recommendation for 1-MNA, its intake or elevation through nicotinamide supplementation may have beneficial effects in treating or delaying cardiac events and decreasing chronic inflammation^15,32^. Spermidine declines with aging^14^, is known to extend lifespan in yeast, nematodes, flies and mice^33^. These benefits likely occur through multiple mechanisms, including stimulating autophagy^14^, reducing cardiac hypertrophy, improving mitochondrial respiration^34^ and reducing the inflammatory response^35^. Similar to spermidine and 1-MNA, PEA extends lifespan and increases survival through anti-inflammatory effects in mouse models^36,37^. PEA also has potential benefits in treating neurodegenerative and CNS disorders^16,38–40^, reducing intracellular lipid accumulation and oxidative stress in mice^41^. Finally, OEA has shown promising results in obesity both in animal models^17,42^ and humans^43,44^. Recent human trials found that 8 weeks of OEA supplementation reduces inflammation (IL-6 and TNF-α) and oxidative stress in obese people^44^. The supplement also reduces weight, BMI, waist circumference, body fat percentage, hunger and cravings, while increasing the expression of fatty acid oxidation modulator proliferator-activated receptor-α (PPAR-α) across a wide range of disease states including PCOS, NAFLD and pre-diabetes^43^. Together, our results support and extend previous animal and human research into the impacts of 1-MNA, spermidine, PEA and OEA on subjective hunger and satiety, as well as oxidative stress and cardiometabolic factors. Importantly, as mentioned above, the data from this study shows that these effects are also not limited to diseased populations and that Mimio can exert similar benefits even in healthy people underscoring its potential for health optimization beyond the resolution of disease symptoms.

The present study was the first randomized, double-blind, placebo-controlled clinical trial to evaluate a fasting mimetic formulation containing nicotinamide, spermidine, PEA, and OEA. Mimio appears safe and well-tolerated and provides benefits to both the subjective experience of hunger and satiety, digestive comfort and objective cardiometabolic blood markers. Importantly, this is also the first study of its kind to show that a portion of the metabolic benefits of prolonged fasting can be phenocopied via simple supplementation with bioactive molecules elevated in the body during prolonged fasting (Mimio). This paves the way for the research and discovery of additional novel fasting mimetic compounds and the development of other biomimetic formulations that may be used to replicate the effects of similar beneficial biochemical states such as exercise. Future study is warranted to establish the longer term impacts of Mimio and evaluation of a larger population of both men and women is necessary to describe potential sex-specific effects. Additionally, given the beneficial impacts on hunger, satiety and digestive comfort, we hypothesize that Mimio may aid in weight loss over a longer period of time and may trigger a new homeostatic setpoint to lipid metabolism. Finally, given the lifespan extending effects demonstrated by Mimio and its constitutive ingredients in previous studies, future studies of Mimio should focus on its hypothesized ability to reduce markers and metrics of biological aging in humans, particularly elderly cohorts for whom fasting would otherwise be unsafe.

## Author Contributions

Study conceptualization: NC, CHR, MCBE, ADG, PLO, AK. Data curation: PLO, ADG. Formal data analysis and visualization: ADG. Funding acquisition: MCBE, NC, CHR. Investigation: MCBE, AK, NC, ADG, JM, VL, PLO. Methodology: NC, MCBE, ADG, PLO, CHR. Project administration: MCBE, AK, PLO, NC, JM, VL. ADG prepared the manuscript with help from MCBE and CHR. All authors edited and approved the final manuscript.

## Acknowledgements

None

## Funding

This study was funded by Mimio Health, LLC

## Competing Interests

Co-author from Mimio had the opportunity to review results generated, but not to eliminate any statistical findings from the report.

## Data Availability

Data are available upon reasonable request from the corresponding author.

## Notes

### Clinical Trial

NCT06790407

## References

1. Longo, V. D. & Mattson, M. P. Fasting: molecular mechanisms and clinical applications. Cell Metab. 19, 181–192 (2014).

2. Mattson, M. P., Longo, V. D. & Harvie, M. Impact of intermittent fasting on health and disease processes. Ageing Res. Rev. 39, 46–58 (2017).

3. Longo, V. D. & Panda, S. Fasting, Circadian Rhythms, and Time-Restricted Feeding in Healthy Lifespan. Cell Metab. 23, 1048–1059 (2016).

4. Lee, C. & Longo, V. Dietary restriction with and without caloric restriction for healthy aging. F1000Research 5, F1000 Faculty Rev-117 (2016).

5. Longo, V. D. & Fontana, L. Calorie restriction and cancer prevention: metabolic and molecular mechanisms. Trends Pharmacol. Sci. 31, 89–98 (2010).

6. Ahmed, A. et al. Impact of intermittent fasting on human health: an extended review of metabolic cascades. Int. J. Food Prop. 21, 2700–2713 (2018).

7. Malinowski, B. et al. Intermittent Fasting in Cardiovascular Disorders—An Overview. Nutrients 11, 673 (2019).

8. Tinsley, G. M. & La Bounty, P. M. Effects of intermittent fasting on body composition and clinical health markers in humans. Nutr. Rev. 73, 661–674 (2015).

9. Azevedo, F. R. D., Ikeoka, D. & Caramelli, B. Effects of intermittent fasting on metabolism in men. Rev. Assoc. Médica Bras. 59, 167–173 (2013).

10. Trepanowski, J. F. et al. Effect of Alternate-Day Fasting on Weight Loss, Weight Maintenance, and Cardioprotection Among Metabolically Healthy Obese Adults: A Randomized Clinical Trial. JAMA Intern. Med. 177, 930–938 (2017).

11. Aoun, A., Ghanem, C., Hamod, N. & Sawaya, S. The Safety and Efficacy of Intermittent Fasting for Weight Loss. Nutr. Today 55, 270 (2020).

12. Laza, V. Intermittent fasting in athletes: PROs and CONs. Health Sports Rehabil. Med. 21, 52–58 (2020).

13. Cienfuegos, S. et al. Effect of Intermittent Fasting on Reproductive Hormone Levels in Females and Males: A Review of Human Trials. Nutrients 14, 2343 (2022).

14. Madeo, F., Bauer, M. A., Carmona-Gutierrez, D. & Kroemer, G. Spermidine: a physiological autophagy inducer acting as an anti-aging vitamin in humans? Autophagy 15, 165–168 (2019).

15. Gebicki, J. et al. 1-Methylnicotinamide: a potent anti-inflammatory agent of vitamin origin. Pol. J. Pharmacol. 55, 109–112 (2003).

16. Mattace Raso, G., Russo, R., Calignano, A. & Meli, R. Palmitoylethanolamide in CNS health and disease. Pharmacol. Res. 86, 32–41 (2014).

17. Thabuis, C., et al. Biological Functions and Metabolism of Oleoylethanolamide. Lipids 43, 887 (2008).

18. Rhodes, C. H. et al. Human fasting modulates macrophage function and upregulates multiple bioactive metabolites that extend lifespan in Caenorhabditis elegans: a pilot clinical study. Am. J. Clin. Nutr. 117, 286–297 (2023).

19. Rhodes, C. H. et al. Absorption, anti-inflammatory, antioxidant, and cardioprotective impacts of a novel fasting mimetic containing spermidine, nicotinamide, palmitoylethanolamide, and oleoylethanolamide: A pilot dose-escalation study in healthy young adult men. Nutr. Res. N. Y. N 132, 125–135 (2024).

20. Stunkard, A. J. & Messick, S. The three-factor eating questionnaire to measure dietary restraint, disinhibition and hunger. J. Psychosom. Res. 29, 71–83 (1985).

21. Ooi, T. C. et al. Intermittent Fasting Enhanced the Cognitive Function in Older Adults with Mild Cognitive Impairment by Inducing Biochemical and Metabolic changes: A 3-Year Progressive Study. Nutrients 12, 2644 (2020).

22. Rast, P., Zimprich, D., Van Boxtel, M. & Jolles, J. Factor Structure and Measurement Invariance of the Cognitive Failures Questionnaire Across the Adult Life Span. Assessment 16, 145–158 (2009).

23. Broadbent, D. E., Cooper, P. F., FitzGerald, P. & Parkes, K. R. The Cognitive Failures Questionnaire (CFQ) and its correlates. Br. J. Clin. Psychol. 21, 1–16 (1982).

24. Karlsson, J., Persson, L. O., Sjöström, L. & Sullivan, M. Psychometric properties and factor structure of the Three-Factor Eating Questionnaire (TFEQ) in obese men and women. Results from the Swedish Obese Subjects (SOS) study. Int. J. Obes. Relat. Metab. Disord. J. Int. Assoc. Study Obes. 24, 1715–1725 (2000).

25. Duran, E. K., et al. Triglyceride-Rich Lipoprotein Cholesterol, Small Dense LDL Cholesterol, and Incident Cardiovascular Disease. J. Am. Coll. Cardiol. 75, 2122–2135 (2020).

26. Jin, X., Yang, S., Lu, J. & Wu, M. Small, Dense Low-Density Lipoprotein-Cholesterol and Atherosclerosis: Relationship and Therapeutic Strategies. Front. Cardiovasc. Med. 8, 804214 (2021).

27. Superko, H. & Garrett, B. Small Dense LDL: Scientific Background, Clinical Relevance, and Recent Evidence Still a Risk Even with ‘Normal’ LDL-C Levels. Biomedicines 10, 829 (2022).

28. Qiao, Y.-N., Zou, Y.-L. & Guo, S.-D. Low-density lipoprotein particles in atherosclerosis. Front. Physiol. 13, (2022).

29. Krauss, R. M. Small Dense LDL Particles: Clinically Relevant? Curr. Opin. Lipidol. 33, 160 (2022).

30. Campos, H., Moye, L. A., Glasser, S. P., Stampfer, M. J. & Sacks, F. M. Low-Density Lipoprotein Size, Pravastatin Treatment, and Coronary Events. JAMA 286, 1468–1474 (2001).

31. Schmeisser, K. et al. Role of Sirtuins in Lifespan Regulation is Linked to Methylation of Nicotinamide. Nat. Chem. Biol. 9, 693–700 (2013).

32. Chlopicki, S. et al. 1-Methylnicotinamide (MNA), a primary metabolite of nicotinamide, exerts anti-thrombotic activity mediated by a cyclooxygenase-2/prostacyclin pathway. Br. J. Pharmacol. 152, 230–239 (2007).

33. Madeo, F., Carmona-Gutierrez, D., Kepp, O. & Kroemer, G. Spermidine delays aging in humans. Aging 10, 2209–2211 (2018).

34. Eisenberg, T. et al. Cardioprotection and lifespan extension by the natural polyamine spermidine. Nat. Med. 22, 1428–1438 (2016).

35. Choi, Y. H. & Park, H. Y. Anti-inflammatory effects of spermidine in lipopolysaccharide-stimulated BV2 microglial cells. J. Biomed. Sci. 19, 31 (2012).

36. Lama, A. et al. Palmitoylethanolamide dampens neuroinflammation and anxiety-like behavior in obese mice. Brain. Behav. Immun. 102, 110–123 (2022).

37. Heide, E. C. et al. Prophylactic Palmitoylethanolamide Prolongs Survival and Decreases Detrimental Inflammation in Aged Mice With Bacterial Meningitis. Front. Immunol. 9, 2671 (2018).

38. Schweiger, V. et al. Ultramicronized Palmitoylethanolamide (um-PEA) as Add-on Treatment in Fibromyalgia Syndrome (FMS): Retrospective Observational Study on 407 Patients. CNS Neurol. Disord. Drug Targets 18, 326–333 (2019).

39. Beggiato, S., Tomasini, M. C. & Ferraro, L. Palmitoylethanolamide (PEA) as a Potential Therapeutic Agent in Alzheimer’s Disease. Front. Pharmacol. 10, 821 (2019).

40. Coppola, M. & Mondola, R. Is there a role for palmitoylethanolamide in the treatment of depression? Med. Hypotheses 82, 507–511 (2014).

41. Annunziata, C. et al. Palmitoylethanolamide counteracts hepatic metabolic inflexibility modulating mitochondrial function and efficiency in diet-induced obese mice. FASEB J. Off. Publ. Fed. Am. Soc. Exp. Biol. 34, 350–364 (2020).

42. Sayd, A. et al. Systemic Administration of Oleoylethanolamide Protects from Neuroinflammation and Anhedonia Induced by LPS in Rats. Int. J. Neuropsychopharmacol. 18, pyu111 (2015).

43. Laleh, P. et al. Oleoylethanolamide increases the expression of PPAR-Α and reduces appetite and body weight in obese people: A clinical trial. Appetite 128, 44–49 (2018).

44. Payahoo, L., Kahjebishak, Y., Jafarabadi, M. A. & Ostadrahimi, A. Oleoylethanolamide Supplementation Reduces Inflammation and Oxidative Stress in Obese People: A Clinical Trial. Adv. Pharm. Bull. 8, 479–487 (2018).

