## Supplemental Figures for "A Novel Fasting Mimetic (Mimio™) Improves Hunger, Digestion, Oxidative Stress, and Cardiometabolic Markers in Overweight Adults with Elevated HbA1c: a Double-Blind, Randomized, Placebo-Controlled Trial"

**Supplemental Materials**

**Supplemental Figure 1. Raw Lab Values Before and After.**

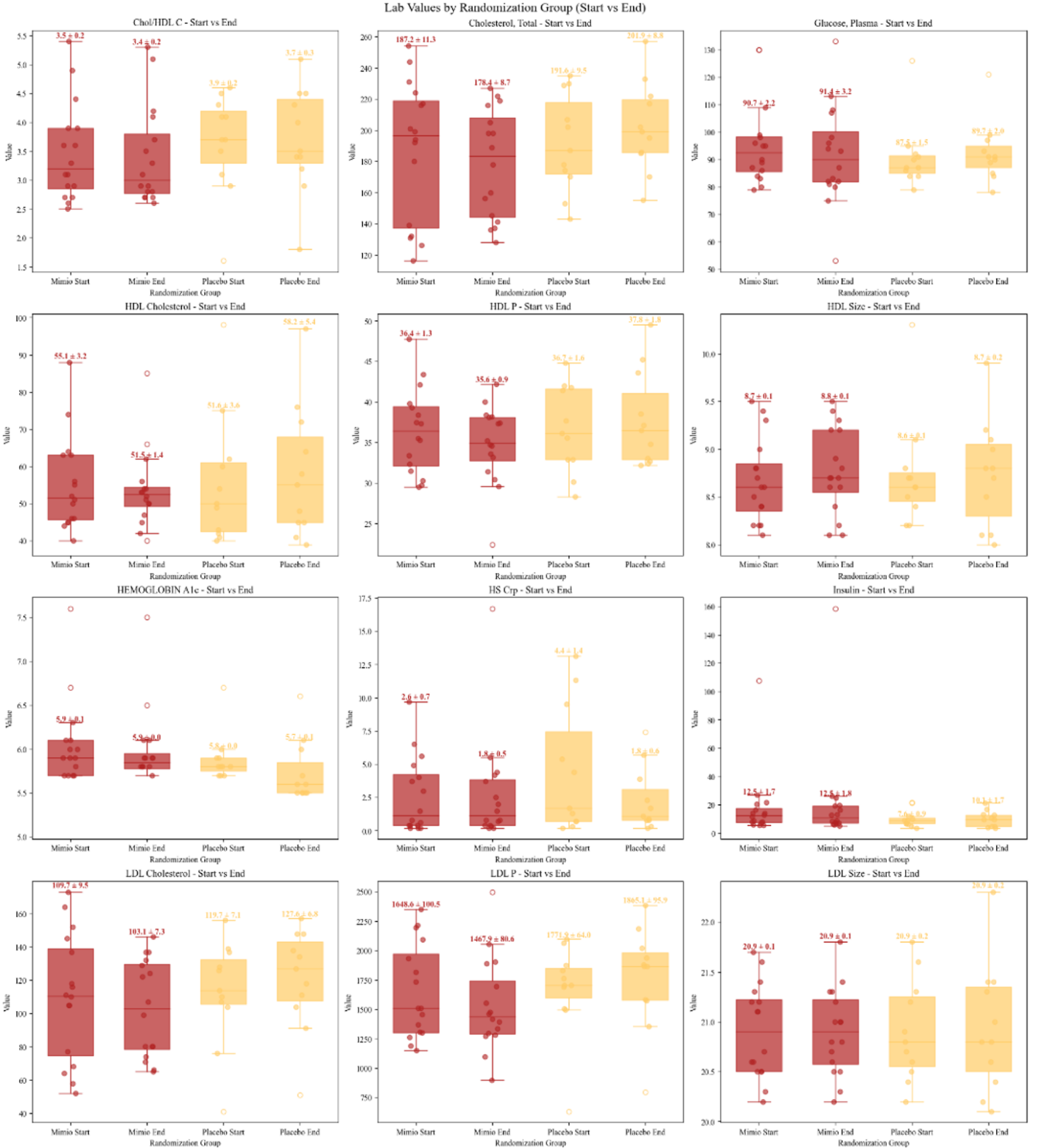

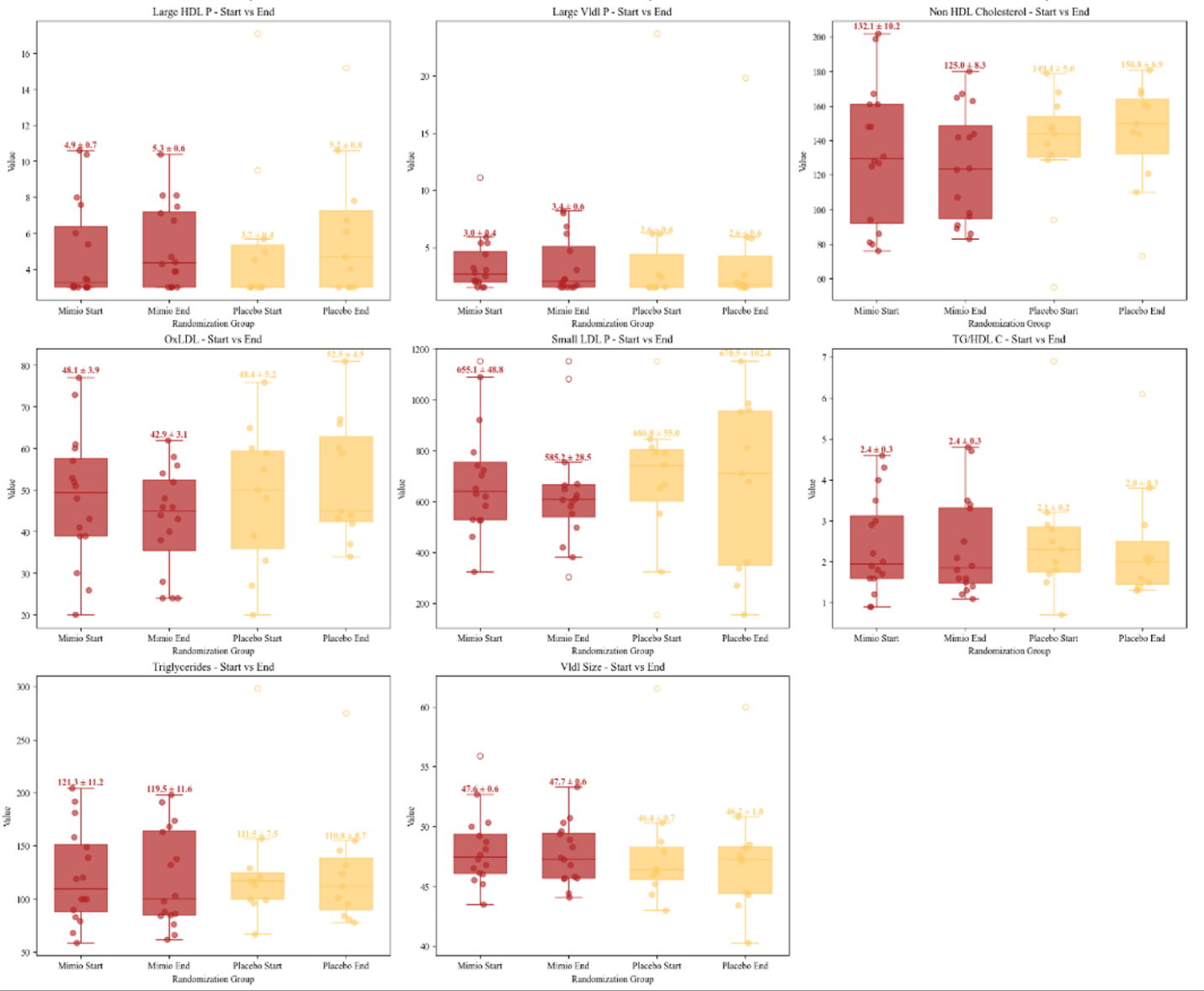

Supplemental Figure 1. Raw Laboratory Blood Values Before and After. Box and whisker plots with mean (SEM) depicted at the top of each whisker for Mimio (red) and placebo (yellow) participants at baseline and end of study (8 weeks). Individual dots represent individual participants’ data. Units can be found in main Table 3.

**Supplemental Table 1. % Improvers in Daily Hunger and Satiety**

| **Daily Metric** | **Mimio**  **% Improved** | **Placebo**  **% Improved** | **p-value** |
| --- | --- | --- | --- |
| **Unhealthy Cravings** | **65** | **73** | **0.74** |
| **Eating Only When Hungry** | **69** | **42** | **0.12** |
| **Distraction From Cravings** | **69** | **63** | **0.75** |
| **Postprandial Satiety** | **60** | **68** | **0.75** |
| **Mealtime Appetite** | **91** | **47** | **0.0025** |
| **Maximum Daily Hunger** | **87** | **58** | **0.04** |
| **Overall Daily Hunger** | **86** | **37** | **0.0011** |
| **Composite Score** | **74** | **53** | **0.2** |

****Metrics with statistically significant differences are shown in bold.***

**Supplemental Figure 2. Weekly Digestive Metrics at Week 8.**

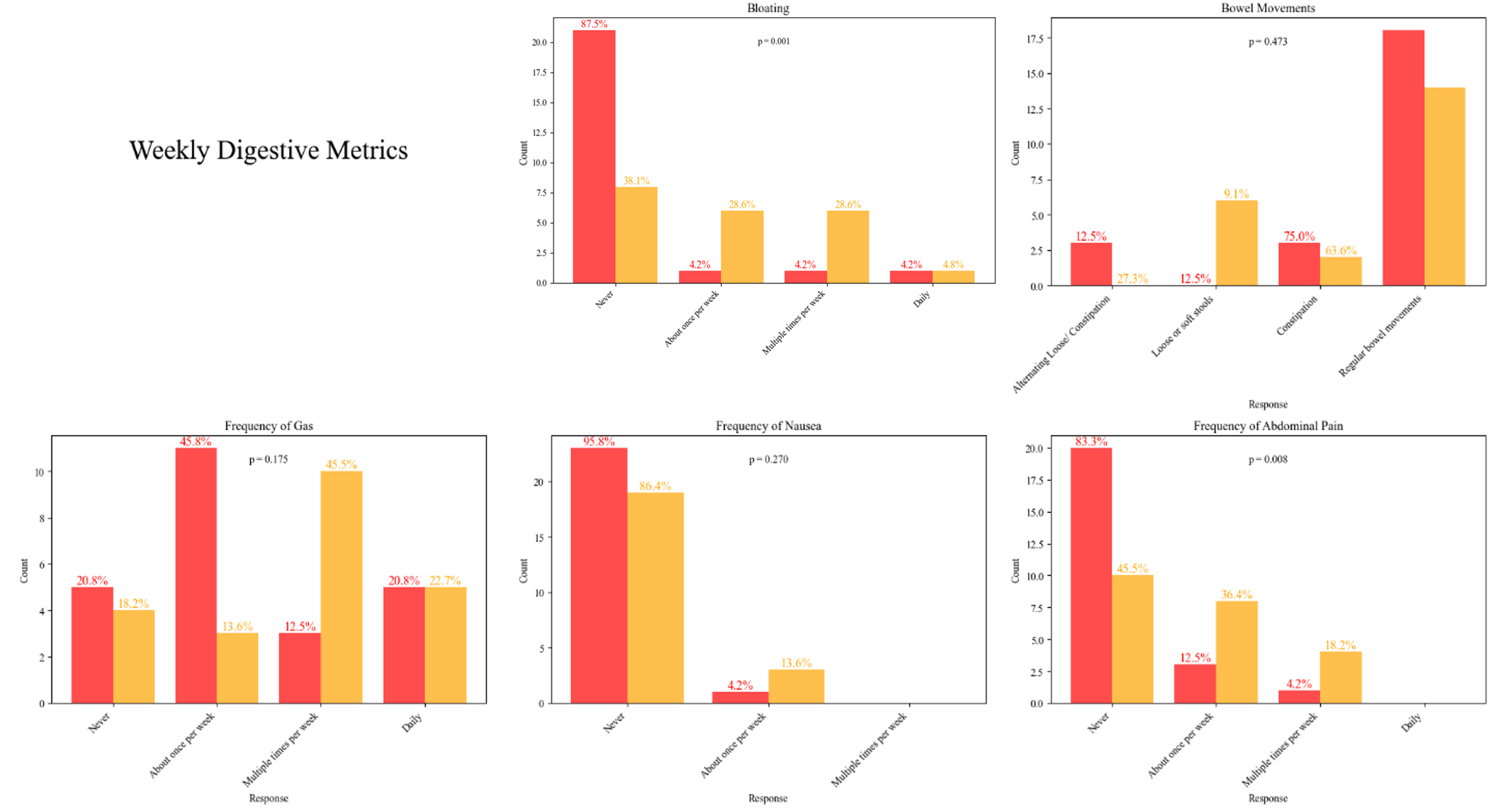

Supplemental Figure 2. Weekly Digestive Metrics at Week 8. Distribution of categorical responses for Mimio (red) and placebo (yellow) participants’ bloating, bowel movement type, frequency of gas, frequency of nausea and frequency of abdominal pain. Percentages represent the percent of that cohort that gave a given response. P-values represent Mann-Whitney U tests for difference in distributions between Mimio and placebo.

**Supplemental Figure 3. Other Weekly Metrics at Week 8**

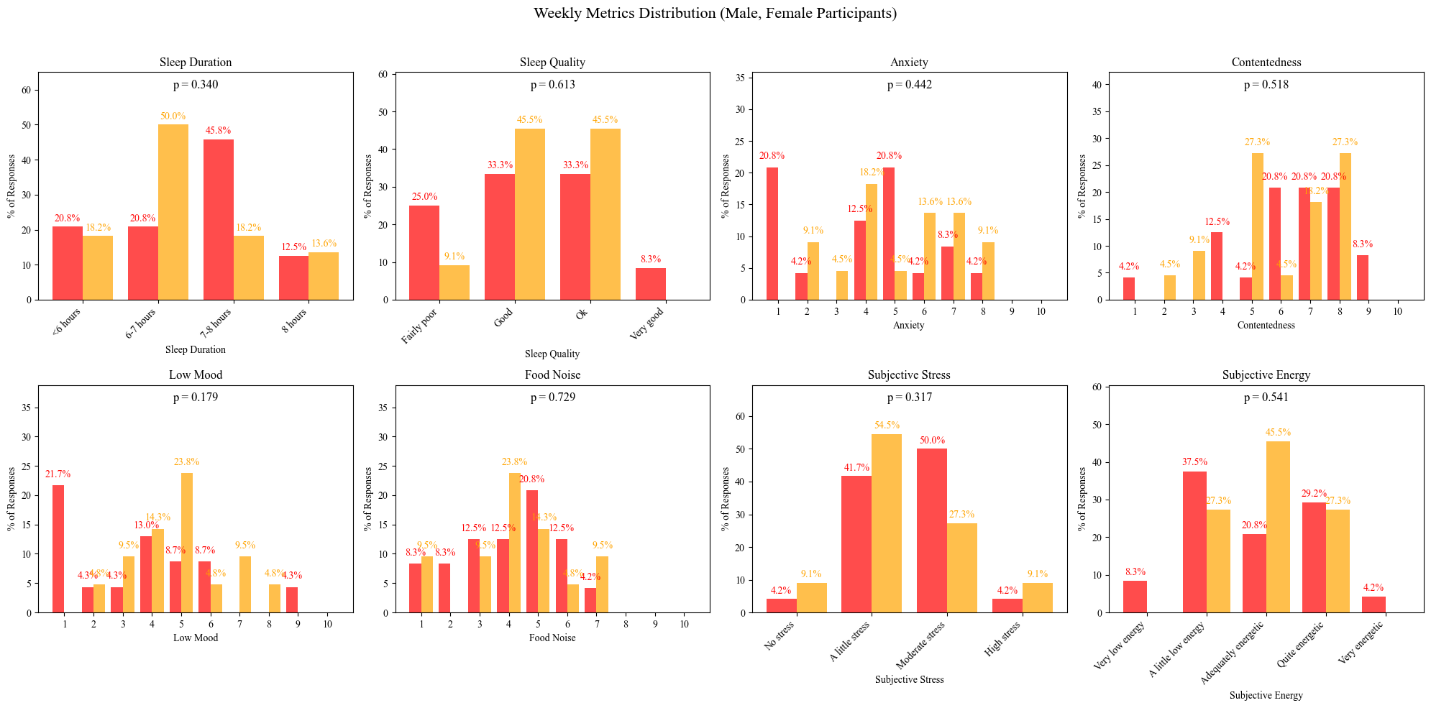

Supplemental Figure 3. Other Weekly Metrics at Week 8. Distribution of categorical responses for Mimio (red) and placebo (yellow) participants’ sleep duration, sleep quality, anxiety, contentedness, low mood, food noise, subjective stress and subjective energy. Percentages represent the percent of that cohort that gave a given response. P-values represent Mann-Whitney U tests for difference in distributions between Mimio and placebo.

**Adverse Events.** 6 AEs occurred during the study, 11 in the Mimio group and 10 in the Placebo group. Only 1 AE, a case of mild diarrhea, occurred during product use in the Mimio group and was considered possibly attributable to the intervention. By comparison, 5 digestive AEs occurred in the Placebo group. Please see Supplemental Table [XX] for all categorized AEs. There were 2 SAEs unrelated to product use (counted under “Other”). One was a case of cellulitis that required hospitalization and IV medication in the Mimio group. The other was a heart attack in the placebo group. 2 additional AEs were categorized as “Other”, and none were attributed to the study intervention. These were a recurrence of leg pain in the Mimio group and a case of lightheadedness in the placebo group.

**Supplemental Table 2. Adverse Events.**

| **AE Type** | **Mimio** | **Placebo** |
| --- | --- | --- |
| **Digestive** | **1** | **5** |
| **Diarrhea** | **1** | **3** |
| **Stomach Pain** | **0** | **2** |
| **Gas** | **0** | **1** |
| **Headache** | **1** | **0** |
| **Cold or Flu** | **2** | **3** |
| **Allergy** | **0** | **1** |
| **Other** | **2** | **2** |

**Supplemental Figure 4. Three Factor Eating Questionnaire (TFEQ-18) Score Change at Week 8.**

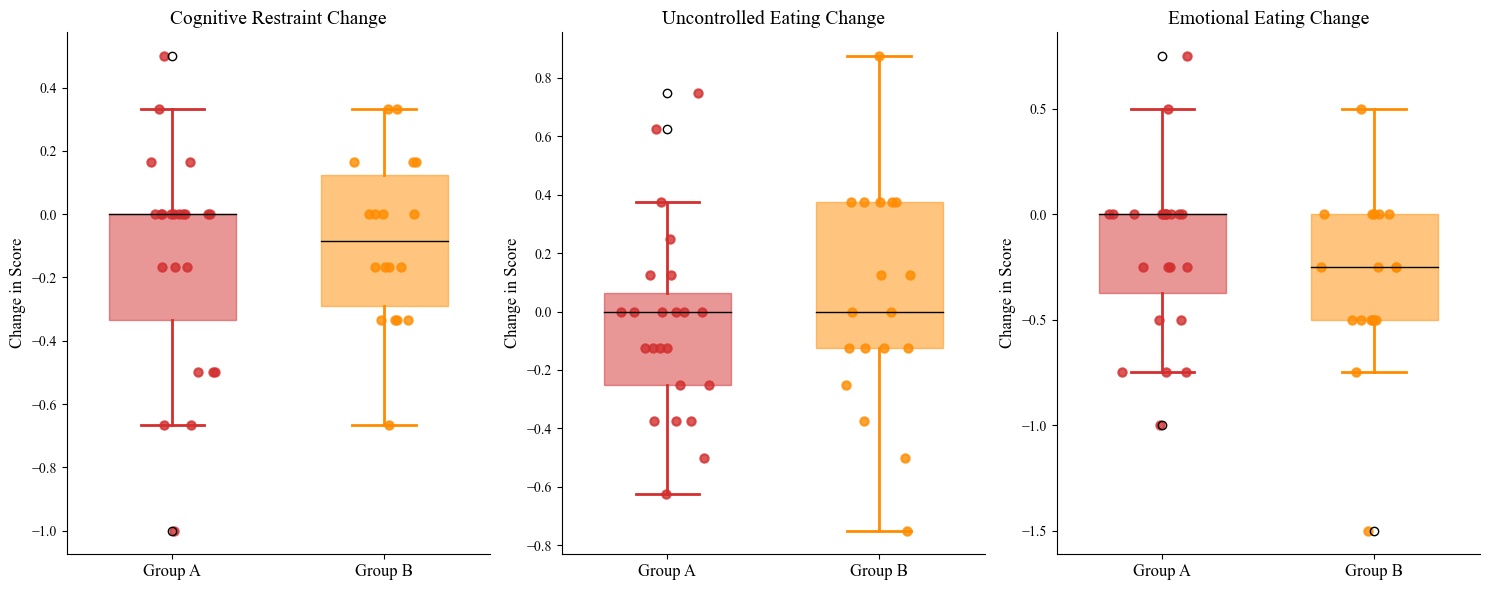

Supplemental Figure 4. Three Factor Eating Questionnaire (TFEQ-18) Score Change at Week 8. Box and whisker plots of change from baseline TFEQ-18 score to week 8 score in Mimio (red) and placebo (yellow) participants. Individual dots represent individual participants’ data.

**Supplemental Figure 5. Three Factor Eating Questionnaire (TFEQ-18) Scores Over Time**

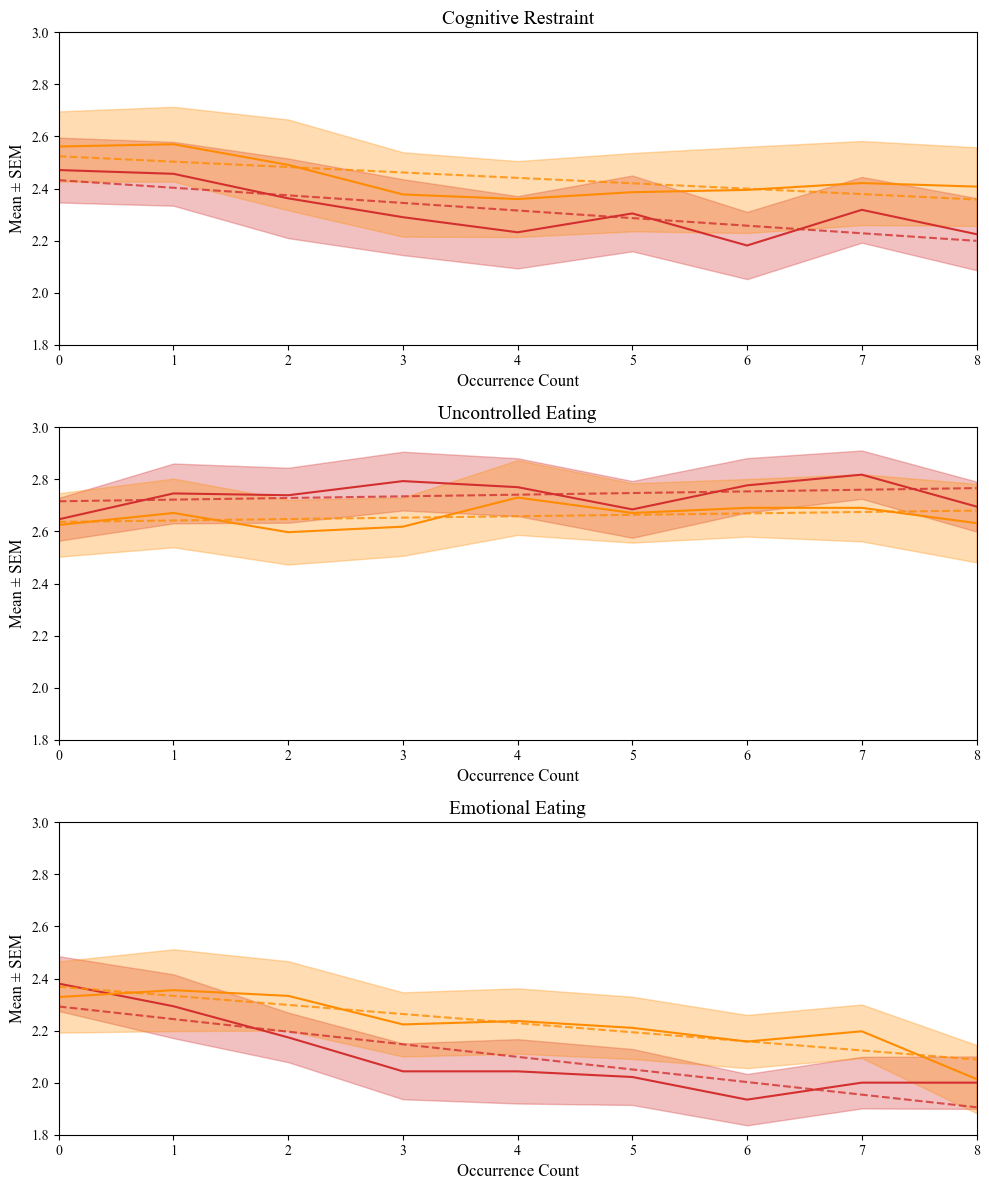

Supplemental Figure 5. TFEQ-18 Scores Over Time. Mean with shaded SEM scores are shown from Week 0 (baseline) through 8 weeks of product or placebo use. Cognitive restraint (top), uncontrolled eating (middle) and emotional eating (bottom) components of the TFEQ-18 are shown separately. Mimio is shown in red and placebo in yellow. Trend lines are graphed for each subcomponent.

**Supplemental Figure 6. Cognitive Failures Questionnaire**

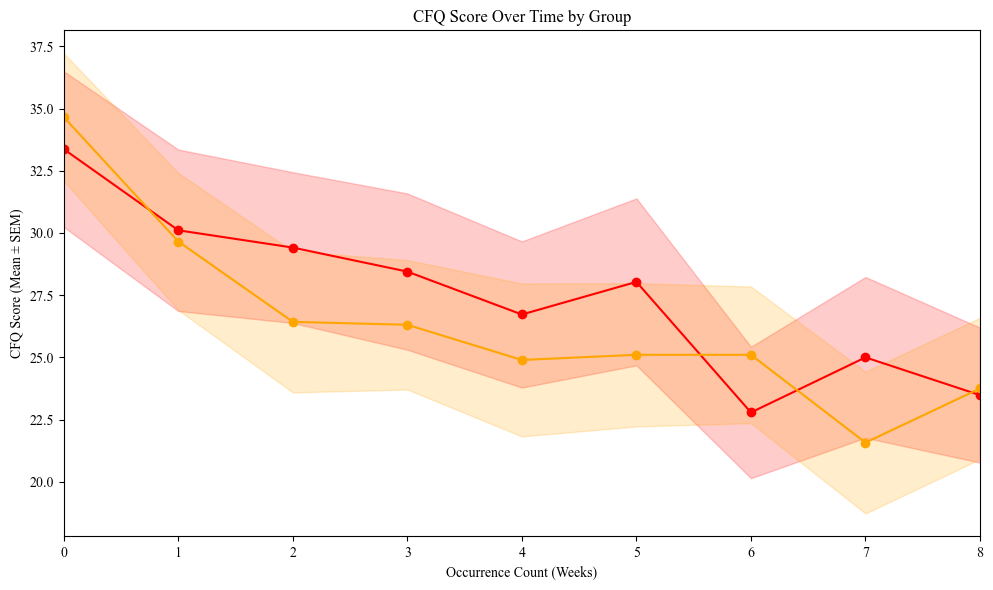

Supplemental Figure 6. Cognitive Failures Questionnaire. Mean with shaded SEM weekly CFQ score in Mimio (red) and placebo (yellow) from baseline (0) to week 8 of Mimio/placebo use.
